# Prognostic Utility of Lactate, Standard Base Excess, and Alactic Base Excess in Sepsis: A Retrospective Analysis of Critical Care Biomarkers

**DOI:** 10.1101/2025.07.29.25332405

**Authors:** Ozgur Kilic, Melda Isevi, Ozkul Yilmaz Colak, Tugcehan Sezer Akman

## Abstract

**Background:** Lactate is an established prognostic marker in sepsis, but the additional predictive value of standard base excess (SBE) and alactic base excess (aBE) remains unclear. This study aimed to evaluate the prognostic utility of lactate, SBE, and aBE in predicting mortality among patients with sepsis and septic shock.

**Methods:** This retrospective cohort study included 218 adult patients admitted to the intensive care unit (ICU) with a diagnosis of sepsis or septic shock. Arterial blood gas parameters (lactate, SBE, and calculated aBE), severity scores (APACHE II, SOFA), and clinical outcomes were recorded. Patients were stratified into survivors and non-survivors. Receiver operating characteristic (ROC) curve analysis and multivariate logistic regression were used to assess the prognostic accuracy of the biomarkers.

**Results:** Among 218 patients, 128 (58.7%) were non-survivors. Non-survivors had significantly higher lactate levels (median: 2.9 mmol/L vs. 1.2 mmol/L; *p* < 0.001). Lactate remained an independent predictor of mortality (OR: 1.40, 95% CI: 1.11–1.77; *p* = 0.005). SBE showed limited prognostic value and lost significance in multivariate analysis. aBE did not differ significantly between groups and was not associated with mortality. ROC analysis showed lactate had the highest area under the curve (AUC: 0.742), while SBE (AUC: 0.421) was a poor predictor.

**Conclusions:** Lactate is a superior independent predictor of ICU mortality in sepsis and septic shock. Neither SBE nor aBE provided additional prognostic value. These findings support the continued use of lactate for risk stratification, while highlighting the limited utility of SBE and aBE in this context.

## Introduction

Sepsis and septic shock remain major global health challenges, characterized by dysregulated host responses to infection and leading to high rates of morbidity and mortality. Early identification of patients at increased risk of adverse outcomes is crucial to guide timely interventions and improve clinical decision-making in intensive care settings. In this context, reliable laboratory biomarkers that reflect underlying pathophysiological disturbances are essential for effective risk stratification [1].

Lactate is among the most widely used and studied biomarkers in sepsis, offering valuable insight into global tissue hypoperfusion, anaerobic metabolism, and mitochondrial dysfunction [2]. Elevated lactate levels have been consistently associated with poor clinical outcomes and have been incorporated into international guidelines for the diagnosis and management of septic shock. However, lactate represents only one component of the complex metabolic derangements that occur during sepsis, and its prognostic value may be influenced by additional acid–base disturbances [3].

Standard base excess (SBE), a calculated parameter derived from arterial blood gas analysis, reflects the net metabolic component of acid–base balance and has been used to quantify the severity of metabolic acidosis [4]. While SBE incorporates the effects of lactate, it is also sensitive to other non-lactate contributors to acidosis, including renal dysfunction, hyperchloremia, and unmeasured anions. The concept of alactic base excess (aBE), defined as SBE plus lactate, was developed to isolate the non-lactate component of metabolic derangement, potentially offering complementary prognostic information in critically ill patients [4].

Despite their physiological relevance, the comparative prognostic performance of lactate, SBE, and aBE in patients with sepsis and septic shock remains poorly defined. Prior studies have yielded conflicting results, and there is limited evidence regarding whether SBE or aBE provide incremental prognostic value beyond lactate alone [5, 6]. Given the widespread availability of these parameters through routine blood gas analysis, their evaluation in a prognostic context holds practical importance.

This study aimed to assess and compare the prognostic accuracy of lactate, SBE, and aBE in predicting in-hospital mortality among patients with sepsis and septic shock. We hypothesized that lactate would remain the most robust prognostic marker and that SBE and aBE would not offer additional predictive value in this population.

## Materials and Methods

### Study Design and Setting

This retrospective observational cohort study was conducted in the medical intensive care unit (ICU) of Ondokuz Mayis University Faculty of Medicine (Samsun, Turkey) between January 2022 and December 2023. The study was approved by the institutional ethics committee (Approval No:2024000579), and the requirement for informed consent was waived due to the retrospective nature of the research.

### Patient Selection

Patients aged ≥18 years who were admitted to the ICU with a diagnosis of sepsis or septic shock, as defined by the Sepsis-3 criteria, were considered eligible. Sepsis was defined as suspected or documented infection accompanied by an acute increase of ≥2 points in the Sequential Organ Failure Assessment (SOFA) score. Septic shock was defined as persistent hypotension requiring vasopressors to maintain a mean arterial pressure (MAP) ≥65 mmHg and serum lactate >2 mmol/L despite adequate fluid resuscitation [7].

Exclusion criteria included: (1) patients with end-stage palliative care, (2) death within 24 hours of ICU admission, and (3) incomplete or missing arterial blood gas (ABG) or laboratory data at ICU admission.

### Data Collection and Laboratory Parameters

Demographic variables, comorbidities (Charlson Comorbidity Index), and clinical severity scores (SOFA and APACHE II) were collected at ICU admission. Arterial blood gas samples were obtained as part of routine clinical assessment within the first hour of ICU admission and were measured using the RAPIDLab 1265 blood gas analyzer (Siemens Healthineers, Erlangen, Germany). The following parameters were recorded: pH, bicarbonate, standard base excess (SBE), and lactate. Alactic base excess (aBE) was calculated as the arithmetic sum of SBE and lactate (aBE = SBE + lactate). Other laboratory data—including serum creatinine, sodium, and chloride levels—were retrieved from the hospital information system. Organ support measures, including vasopressor therapy, mechanical ventilation, and renal replacement therapy, were documented. ICU length of stay and in-hospital mortality were recorded as outcomes.

### Statistical Analysis

Data were analyzed using SPSS software (version 22.0; IBM Corp., Armonk, NY). Continuous variables were assessed for normality using the Shapiro-Wilk test. Normally distributed variables were expressed as mean ± standard deviation and compared using the independent samples t-test. Non-normally distributed variables were reported as median (interquartile range) and compared using the Mann–Whitney U test. Categorical variables were expressed as frequency (%) and analyzed using the chi-square test.

To assess the prognostic performance of lactate, SBE, and aBE in predicting mortality, receiver operating characteristic (ROC) curve analysis was performed, and the area under the curve (AUC) was calculated. The Youden Index was used to determine optimal cutoff values. Variables with a *p* < 0.10 in univariate analysis were included in multivariate logistic regression models to identify independent predictors of mortality. Results were reported as odds ratios (OR) with 95% confidence intervals (CI). A two-tailed *p*-value < 0.05 was considered statistically significant.

Missing data were handled using multiple imputation when the proportion of missing values was <5%; variables with >5% missingness were excluded from the analysis.

## Results

### Patient Characteristics

A total of 218 patients met the inclusion criteria, of whom 90 (41.3%) survived and 128 (58.7%) died during hospitalization. There were no significant differences in age (mean 66.7 ± 15.2 years vs. 66.6 ± 13.9 years, *p* = 0.990) or sex distribution (male 56.3% vs. 65.6%, *p* = 0.074) between survivors and non-survivors.

Non-survivors had significantly higher median APACHE II scores (28 [23–33] vs. 23 [18–27], *p* < 0.001) and SOFA scores (12 [9–14] vs. 9 [7–12], *p* < 0.001). Comorbidity burden, as assessed by the Charlson Comorbidity Index, did not significantly differ between groups (*p* = 0.308). Table 1 shows the demographic and clinical characteristics of survivors and non-survivors, including their age, sex, severity scores (APACHE II, SOFA), and other relevant biomarkers.

**Table 1.**
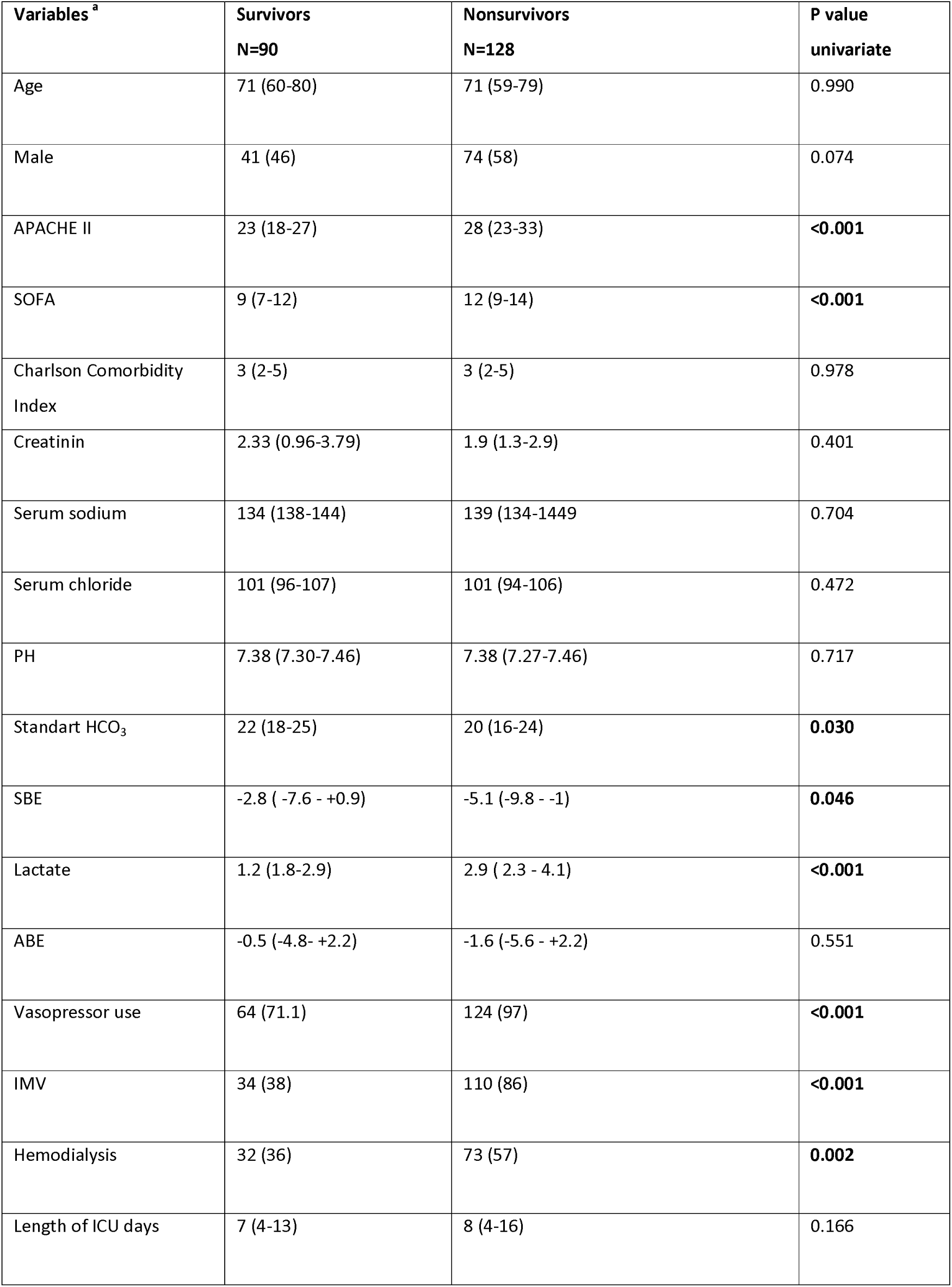
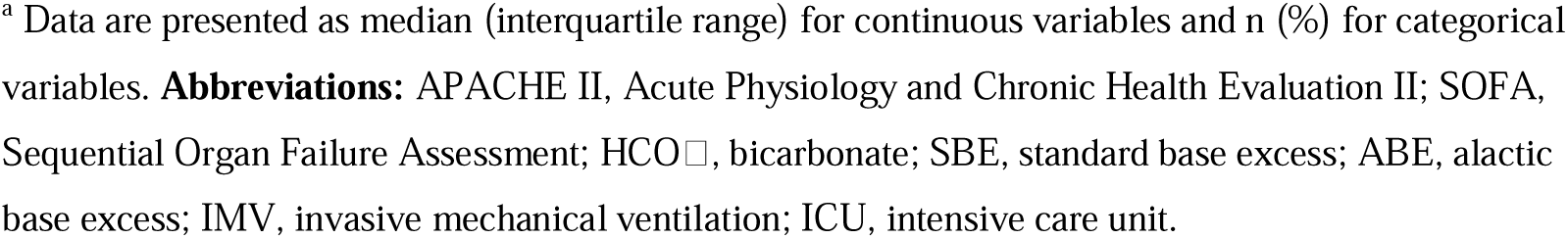
Demographic and clinical characteristics between survivors and non-survivors in patients with sepsis and septic shock.

### Arterial Blood Gas and Laboratory Findings

Key arterial blood gas variables are summarized in Table 1. Lactate concentrations were significantly elevated in non-survivors compared to survivors (2.9 mmol/L [2.3–4.1] vs. 1.9 mmol/L [1.2–2.9], p < 0.001). SBE values were more negative in non-survivors (–5.1 mmol/L [–9.8 to –1.0] vs. –2.8 mmol/L [–7.6 to +0.9], p = 0.046), while aBE did not significantly differ between groups (–1.6 mmol/L [–5.6 to +2.2] vs. –0.5 mmol/L [–4.8 to +2.2], p = 0.551). Bicarbonate levels were lower in non-survivors (20 mmol/L [16–24] vs. 22 mmol/L [18–25], p = 0.030).

### Multivariate Predictors of Mortality

Variables with p < 0.10 in univariate comparisons were entered into a multivariate logistic regression model. Elevated lactate levels (OR: 1.40 per 1 mmol/L increase; 95% CI: 1.11–1.77; p = 0.005), APACHE II score (OR: 1.08 per point; 95% CI: 1.02–1.15; p = 0.013), vasopressor use (OR: 1.32; 95% CI: 1.05–1.68; p = 0.028), and invasive mechanical ventilation (OR: 1.45; 95% CI: 1.10–1.92; p = 0.012) remained independently associated with mortality. Neither SBE nor aBE retained significance in multivariate analysis. Table 2 presents the multivariate logistic regression analysis for mortality, highlighting the significant predictors.

**Table 2.** Multivariate Logistic Regression Analysis for Mortality.

| Variables | OR | %95 CI | P value |
| --- | --- | --- | --- |
| APACHE II score | 1.08 | 1.02-1.15 | 0.013 |
| Lactate level | 1.40 | 1.11-1.77 | 0.005 |
| Vasopressore use | 1.32 | 1.05-1.68 | 0.028 |
| IMV | 1.45 | 1.10-1.92 | 0.012 |

**Abbreviations:** APACHE II; Acute Physiology and Chronic Health Evaluation II, OR; Odds Ratio, CI; Confidence Interval, IMV; Invasive Mechanical Ventilation

### Receiver Operating Characteristic (ROC) Analysis

ROC curve analysis demonstrated that lactate had the highest prognostic accuracy among the tested variables, with an AUC of 0.742 (95% CI: 0.672–0.812; p < 0.001) (see Figure 1). The optimal cutoff value for lactate was 2.0 mmol/L, yielding a sensitivity of 86.7% and specificity of 60.0%. SBE exhibited poor discriminatory ability (AUC: 0.421; 95% CI: 0.343–0.498; p = 0.046). ROC analysis was not performed for aBE due to lack of statistical association in univariate analysis. Figure 2 shows box-and-whisker plots comparing lactate and SBE levels in survivors versus non-survivors.

**Figure 1.**
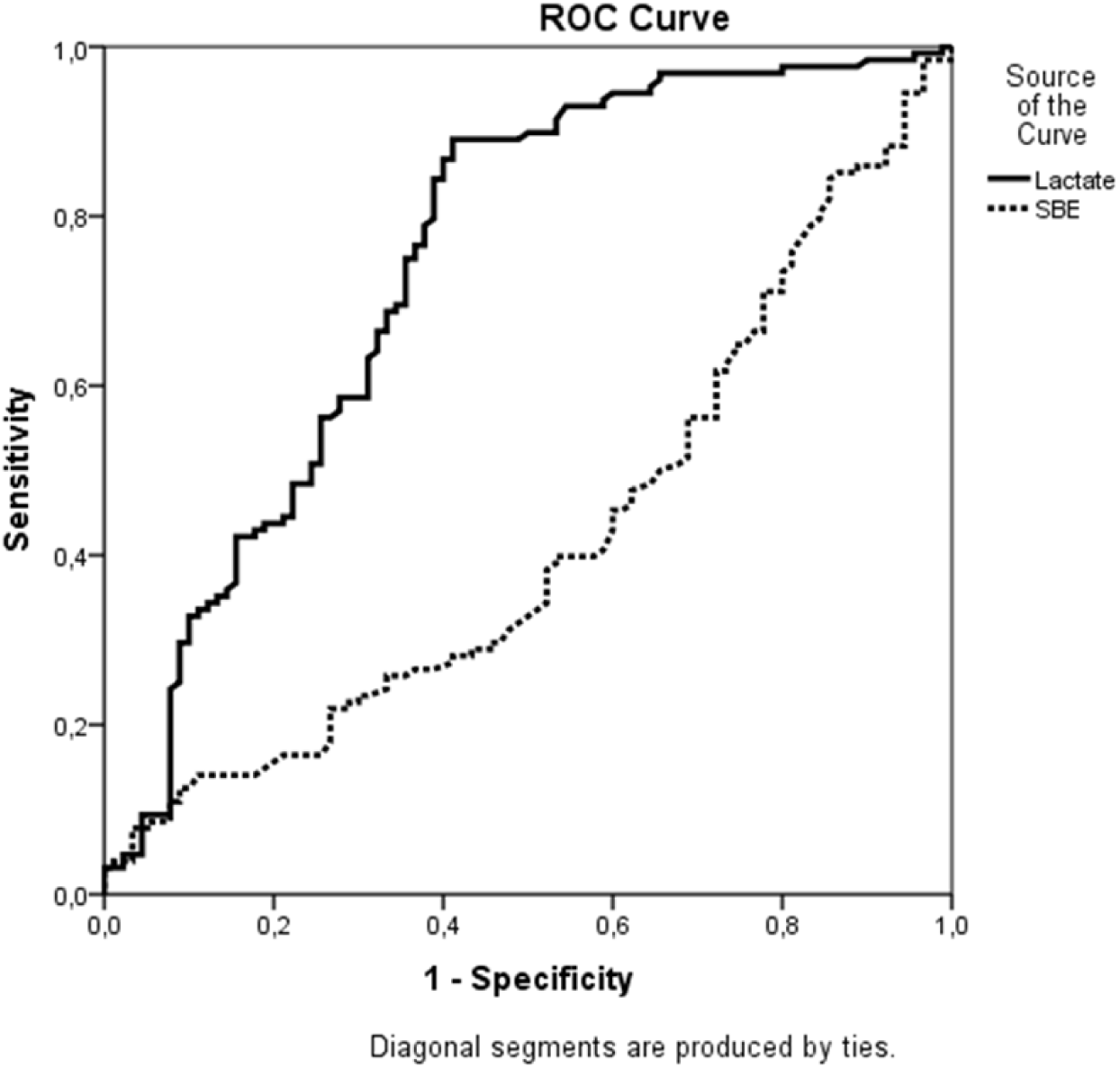
ROC Curve for Lactate and Standard Base Excess. ROC curve demonstrating the prognostic accuracy of lactate and SBE in predicting in-hospital mortality. Lactate had an AUC of 0.742, SBE had an AUC of 0.421.

**Figure 2.**
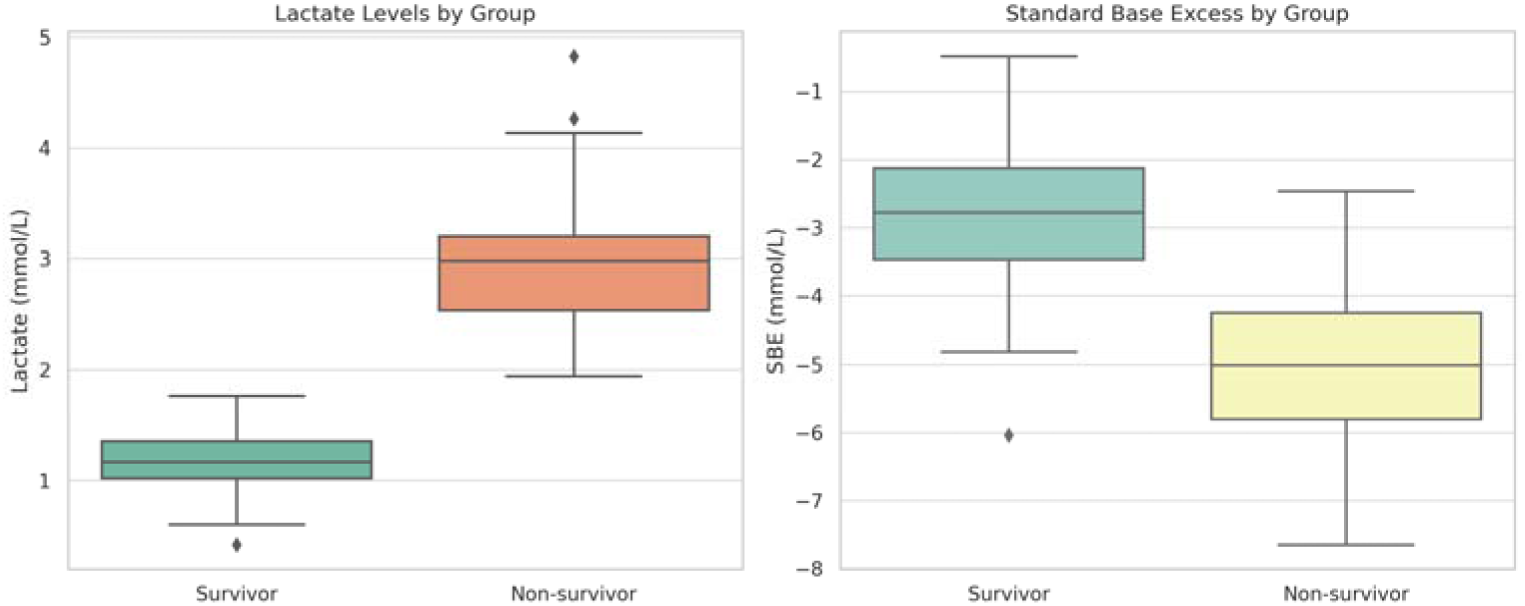
Boxplots of Lactate and Standard Base Excess by Survival Status. Box-and-whisker plots comparing lactate and SBE levels in survivors vs. non-survivors.

### Clinical Interventions and Outcomes

Non-survivors required significantly more intensive interventions: vasopressors (97% vs. 71.1%, p < 0.001), invasive mechanical ventilation (86% vs. 38%, p < 0.001), and renal replacement therapy (57% vs. 36%, p = 0.002). Median ICU length of stay did not differ significantly between groups (8 days [4–16] vs. 7 days [4–13], p = 0.166). These findings are visually represented in Figure 3, which shows a bar chart illustrating the use of vasopressors, mechanical ventilation, and renal replacement therapy by survival status.

**Figure 3.**
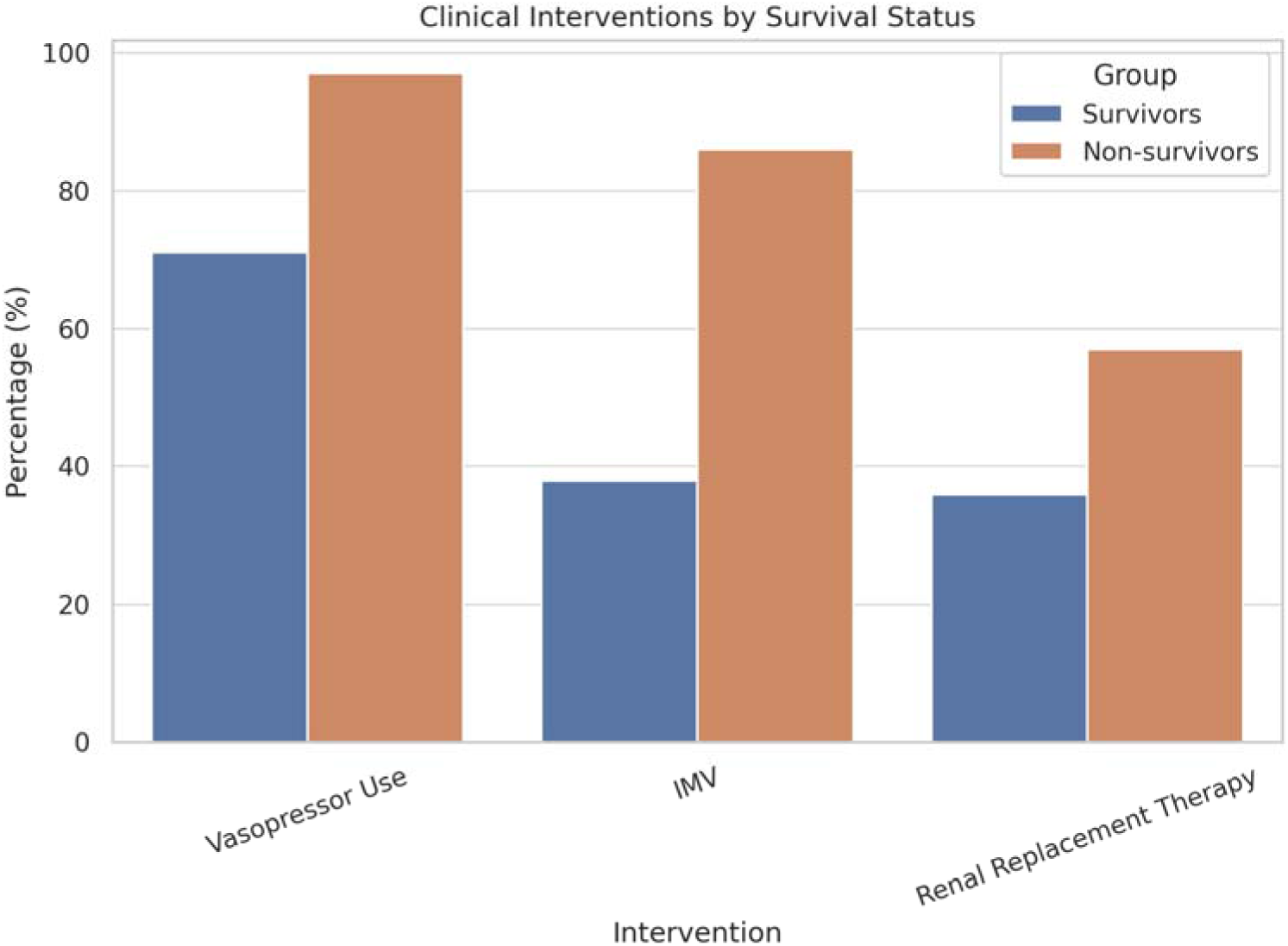
Bar Chart of Clinical Interventions by Survival Group. Bar chart illustrating vasopressor use, mechanical ventilation, and renal replacement therapy by survival status.

### Alactic Base Excess (aBE) Analysis

Additionally, we analyzed aBE, which theoretically isolates non-lactate metabolic disturbances by excluding lactate from the calculation. However, aBE did not show a significant association with mortality. Figure 4 displays the boxplot of aBE values in survivors and non-survivors. No significant difference was observed (p > 0.05).

**Figure 4.**
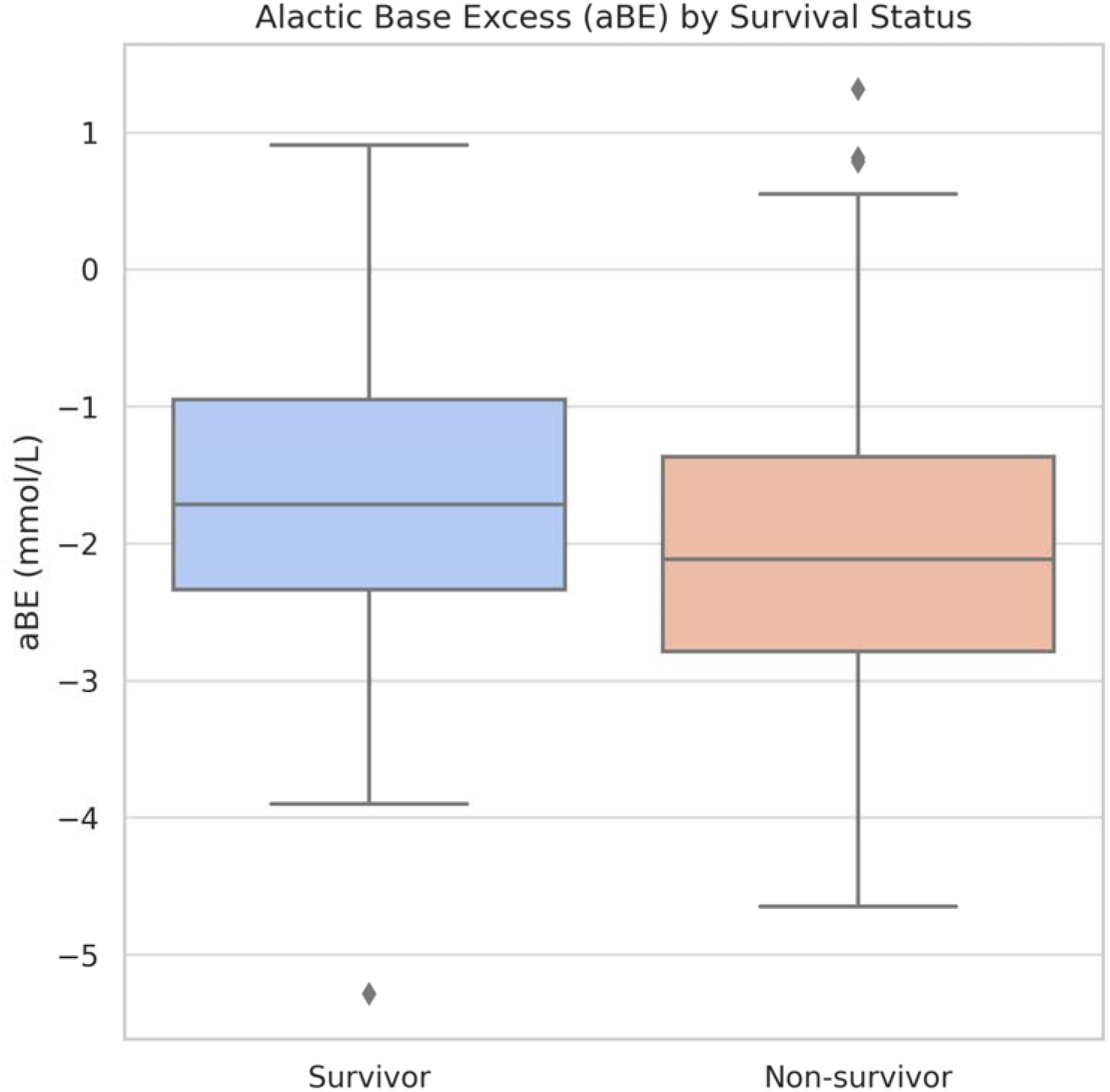
Boxplot of Alactic Base Excess by Survival Status. Boxplot of aBE values in survivors and non-survivors. No significant difference was observed (p > 0.05).

## Discussion

In this study, we investigated the prognostic value of lactate, standard base excess (SBE), and alactic base excess (aBE) in predicting mortality among patients with sepsis and septic shock. Our findings indicate that lactate is a strong and independent predictor of mortality, whereas SBE and aBE do not provide additional prognostic value. These results have significant implications for risk stratification in sepsis and the use of metabolic biomarkers in clinical practice.

Unlike previous studies, our findings align with existing evidence supporting lactate as a critical biomarker in sepsis prognosis [8–10]. Research has shown that lactate levels above 2 mmol/L are associated with significantly increased hospital mortality, even in hemodynamically stable sepsis patients [11]. Additionally, meta-analyses have suggested that changes in lactate levels over time may be more predictive of mortality than a single measurement [12, 13]. However, unlike studies evaluating lactate kinetics, this analysis focuses solely on the initial lactate value at ICU admission without assessing variations over time.

Lactate is a well-known biomarker of impaired oxygen utilization, mitochondrial dysfunction, and anaerobic metabolism [14]. Among the patients analyzed, non-survivors had significantly higher lactate levels than survivors, with this difference being statistically significant. The receiver operating characteristic analysis confirmed the strong predictive performance of lactate, with an optimal cutoff of 2.0 mmol/L, providing high sensitivity (86.7%) and acceptable specificity (60.0%). These findings reinforce the importance of early lactate monitoring in sepsis management and suggest that dynamic changes in lactate levels over time may further improve risk assessment.

SBE quantifies the amount of acid or base required to return blood pH to 7.4 under standard conditions, making it a widely used parameter for evaluating metabolic acidosis and overall acid-base balance [4, 15]. Several studies have highlighted the clinical relevance of SBE in critically ill patients, particularly in sepsis and trauma, where metabolic acidosis often plays a crucial role in disease progression and outcomes [16, 17].

In our analysis, while SBE was significantly associated with mortality in univariate analysis, this association was not maintained in multivariate models, suggesting that its prognostic value is largely overshadowed by lactate. This finding aligns with previous research indicating that lactate, as a marker of impaired tissue oxygenation and anaerobic metabolism, often outperforms SBE in predicting mortality in critically ill patients [8, 10]. The stronger predictive role of lactate may stem from its direct reflection of tissue hypoxia and mitochondrial dysfunction, whereas SBE represents a more generalized measure of acid-base balance influenced by multiple metabolic factors.

Additionally, we analyzed aBE, which theoretically isolates non-lactate metabolic disturbances by excluding lactate from the calculation. However, aBE did not show a significant association with mortality. This finding is consistent with prior studies suggesting that while lactate-independent metabolic acidosis contributes to disease severity, its prognostic impact is less pronounced when compared to lactate-driven acidosis [18, 19]. Given these results, lactate remains the dominant metabolic marker for risk stratification in critically ill patients, reinforcing the importance of dynamic lactate monitoring in clinical practice.

Several factors may explain the limited prognostic utility of SBE and aBE in sepsis. First, metabolic acidosis in sepsis is primarily driven by lactate accumulation. Since lactate is already a strong prognostic marker, SBE, which inherently incorporates lactate-driven acidosis, may not provide additional independent prognostic value. aBE, which aims to differentiate non-lactate acidosis, may be less relevant in sepsis, where lactate is the dominant driver of metabolic acidosis.

Another possible explanation for the lack of prognostic value in SBE and aBE is the influence of various compensatory mechanisms. Unlike lactate, which directly reflects tissue perfusion abnormalities and anaerobic metabolism, SBE and aBE are affected by multiple physiological adaptations, including renal bicarbonate regulation, respiratory compensation, and electrolyte disturbances [20]. For example, patients with sepsis-induced metabolic acidosis may develop compensatory respiratory alkalosis through hyperventilation, normalizing SBE values despite the severity of their condition. Similarly, renal compensation may buffer metabolic acidosis over time, modifying SBE and aBE values independently of sepsis severity [21]. These physiological adaptations likely dilute the prognostic significance of SBE and aBE, explaining their weaker association with mortality compared to lactate.

Previous research on the prognostic value of SBE and aBE has yielded inconsistent results [22–25]. Some findings suggest that more negative SBE values, indicating severe metabolic acidosis, are associated with increased mortality in critically ill patients. However, differences in patient populations across studies complicate direct comparisons. While some analyses have identified SBE as a predictor of mortality in general ICU settings, its prognostic significance appears to diminish in sepsis-specific cases [22]. Similarly, base excess has shown relevance in trauma-related outcomes but lacks the same predictive strength in sepsis [25]. The present analysis contributes to this discussion by suggesting that SBE and aBE may not serve as reliable independent prognostic markers in sepsis, where lactate remains the dominant indicator. Further investigation is required to determine whether these parameters hold predictive value in specific subgroups, such as patients with concurrent renal failure or electrolyte imbalances.

One of the key strengths of our study is its focused comparison of three metabolic biomarkers within a well-defined cohort of sepsis and septic shock patients. The statistical analyses were robust, and the study design allowed for meaningful comparisons between survivors and non-survivors. However, our study also has several limitations.

First, its retrospective and single-center design may limit the generalizability of our findings. Additionally, we only assessed biomarker levels at ICU admission, without considering dynamic changes over time. Sepsis is a rapidly evolving condition, and serial biomarker measurements may provide better prognostic insights than a single baseline value. Future studies should investigate how trends in lactate, SBE, and aBE correlate with patient outcomes. Furthermore, we did not analyze the impact of sepsis etiology or coexisting conditions on biomarker performance. Since different infection sources and comorbidities can influence metabolic responses, future research should explore whether certain patient subgroups may benefit from SBE- or aBE-based prognostic assessment.

## Conclusion

This study confirms that among routinely available metabolic biomarkers, lactate is the most reliable early predictor of mortality in sepsis and septic shock. Neither standard base excess nor alactic base excess provided independent prognostic value. These findings reinforce the clinical utility of lactate-guided risk assessment and suggest caution in interpreting derived acid–base variables without robust validation. Prospective studies are needed to evaluate the role of dynamic biomarker trends and to determine whether specific patient subgroups may benefit from additional acid–base stratification.

## Author Contributions

O. Kilic and M. Isevi contributed to study conception and design. O. Kilic performed data analysis and interpretation. M. Isevi, O.Y. Colak, and T.S. Akman were involved in data acquisition and clinical oversight. The manuscript was drafted by O. Kilic and critically revised by all authors. All authors read and approved the final manuscript.

## Ethics Approval and Consent to Participate

The study protocol was approved by the Ethics Committee of Ondokuz Mayis University Faculty of Medicine, Samsun, Turkey (Approval No: 2024000579). Given the retrospective nature of the study, the need for informed consent was waived.

## Conflict of Interest

The authors declare no conflicts of interest.

## Funding

This research did not receive any specific grant from funding agencies in the public, commercial, or not-for-profit sectors.

## Data Availability

The datasets generated and analyzed during the current study are available from the corresponding author on reasonable request.

## Notes

### Competing Interest Statement

The authors have declared no competing interest.

### Funding Statement

This study did not receive any funding

### Author Declarations

The study protocol was approved by the Ethics Committee of Ondokuz Mayis University Faculty of Medicine, Samsun, Turkey (Approval No: 2024000579).

## References

1. Mukherjee S, Das S, Mukherjee S, Ghosh PS, Bhattacharya S. Arterial blood gas as a prognostic indicator in patients with sepsis. Indian J Med Microbiol. 2020;38(3-4):457–460. doi:10.4103/ijmm.IJMM_19_478.

2. De Backer D, Vincent JL. Understanding hyperlactatemia in human sepsis: Are we making progress? Am J Respir Crit Care Med. 2019;200(8):1070–1071. doi:10.1164/rccm.201905-1024LE.

3. Yang K, Fan M, Wang X, et al. Lactate induces vascular permeability via disruption of VE-cadherin in endothelial cells during sepsis. Sci Adv. 2022;8(17):eabm8965. doi:10.1126/sciadv.abm8965.

4. Gattinoni L, Vasques F, Camporota L, et al. Understanding lactatemia in human sepsis: Potential impact for early management. Am J Respir Crit Care Med. 2019;200(5):582–589. doi:10.1164/rccm.201812-2342OC.

5. Cantos J, Huespe IA, Sinner JF, et al. Alactic base excess is an independent predictor of death in sepsis: A propensity score analysis. J Crit Care. 2023;74:154248. doi:10.1016/j.jcrc.2022.154248.

6. Smuszkiewicz P, Jawień N, Szrama J, Lubarska M, Kusza K, Guzik P. Admission lactate concentration, base excess, and alactic base excess predict the 28-day inward mortality in shock patients. J Clin Med. 2022;11(20):6125. doi:10.3390/jcm11206125.

7. Singer M, Deutschman CS, Seymour CW, et al. The Third International Consensus Definitions for Sepsis and Septic Shock (Sepsis-3). JAMA. 2016;315(8):801–810. doi:10.1001/jama.2016.0287.

8. Jansen TC, van Bommel J, Schoonderbeek FJ, et al. Early lactate-guided therapy in intensive care unit patients: a multicenter, open-label, randomized controlled trial. Am J Respir Crit Care Med. 2010;182(6):752–761. doi:10.1164/rccm.200912-1918OC.

9. Shapiro NI, Howell MD, Talmor D, et al. Serum lactate as a predictor of mortality in emergency department patients with infection. Ann Emerg Med. 2005;45(5):524–528. doi:10.1016/j.annemergmed.2004.12.006.

10. Mikkelsen ME, Miltiades AN, Gaieski DF, et al. Serum lactate is associated with mortality in severe sepsis independent of organ failure and shock. Crit Care Med. 2009;37(5):1670–1677. doi:10.1097/CCM.0b013e31819fcf68.

11. Vincent JL, Quintairos e Silva A, Couto L, et al. The value of blood lactate kinetics in critically ill patients: a systematic review. Crit Care. 2016;20:257. doi:10.1186/s13054-016-1403-5.

12. Zhang Z, Xu X. Lactate clearance is a useful biomarker for the prediction of all-cause mortality in critically ill patients: a systematic review and meta-analysis. Crit Care Med. 2014;42(9):2118–2125. doi:10.1097/CCM.0000000000000405.

13. Haas SA, Lange T, Saugel B, et al. Severe hyperlactatemia, lactate clearance and mortality in unselected critically ill patients. Intensive Care Med. 2016;42(2):202–210. doi:10.1007/s00134-015-4127-0.

14. Gladden LB. Lactate metabolism: a new paradigm for the third millennium. J Physiol. 2004;558(1):5–30. doi:10.1113/jphysiol.2003.058701.

15. Kellum JA. Determinants of blood pH in health and disease. Crit Care. 2000;4(1):6–14. doi:10.1186/cc644.

16. Noritomi DT, Soriano FG, Kellum JA, et al. Metabolic acidosis in patients with severe sepsis and septic shock: a longitudinal quantitative study. Crit Care Med. 2009;37(10):2733–2739. doi:10.1097/ccm.0b013e3181a59165.

17. Vincent JL, Moreno R. Clinical review: scoring systems in the critically ill. Crit Care. 2010;14(2):207. doi:10.1186/cc8204.

18. Andersen LW, Mackenhauer J, Roberts JC, et al. Etiology and therapeutic approach to elevated lactate levels. Mayo Clin Proc. 2013;88(10):1127–1140. doi:10.1016/j.mayocp.2013.06.012.

19. Al-Jaghbeer M, Kellum JA. Acid-base disturbances in intensive care patients: etiology, pathophysiology, and treatment. Nephrol Dial Transplant. 2015;30(7):1104–1111. doi:10.1093/ndt/gfu289.

20. Seifter JL, Chang HY. Disorders of acid-base balance: new perspectives. Kidney Dis [Basel]. 2017;2(4):170–186. doi:10.1159/000453028.

21. Dubin A, Menises MM, Masevicius FD, et al. Comparison of three different methods of evaluation of metabolic acid-base disorders. Crit Care Med. 2007;35(5):1264–1270. doi:10.1097/01.CCM.0000259536.11943.90.

22. Cusack RJ, Rhodes A, Lochhead P, et al. The strong ion gap does not have prognostic value in critically ill patients in a mixed medical/surgical adult ICU. Intensive Care Med. 2002;28(7):864–869. doi:10.1007/s00134-002-1318-2.

23. Park M, Maciel AT, Noritomi DT, et al. Effect of PaCOD variation on standard base excess value in critically ill patients. J Crit Care. 2009;24(4):484–491. doi:10.1016/j.jcrc.2008.12.018.

24. Park M, Taniguchi LU, Noritomi DT, et al. Clinical utility of standard base excess in the diagnosis and interpretation of metabolic acidosis in critically ill patients. Braz J Med Biol Res. 2008;41(3):241–249. doi:10.1590/s0100-879x2006005000199.

25. Diao MY, Wang T, Cui YL, Lin ZF. Zhonghua Wei Zhong Bing Ji Jiu Yi Xue. 2013;25(4):211–214. doi:10.3760/cma.j.issn.2095-4352.2013.04.008.

